# Classifying Texas counties using ARIMA Models on COVID-19 daily confirmed cases: the impact of political affiliation and face covering orders

**DOI:** 10.1101/2021.06.02.21258221

**Authors:** Hao Jiang, Brigitta Pulins, Aurélie C. Thiele

## Abstract

The aim of this paper is to investigate whether the 254 Texas counties in the United States can be grouped in a meaningful way according to the characteristics of the ARIMA or seasonal ARIMA models fitting the logarithm of daily confirmed cases of the Coronavirus Disease 2019 (COVID-19) for 254 counties in Texas of the United States. We analyze clusters of the model’s non-seasonal parameters (*p, d, q*), distinguishing between county-level political affiliations and face covering orders, and also consider county-level population and poverty rate. Using data from March 4, 2020 to March 15, 2021, we find that 223 of the total 254 counties are clustered into 23 model parameters (*p, d, q*), while the number of cases in the remaining 31 counties could not be successfully fitted to ARIMA models. We also find the impact of the county-level infection rate and the county-level poverty rate on clusters of counties with different political affiliations and face covering orders. Further, we find that the infection rate and the poverty rate had a significant high positive correlation, and Democrat-leaning counties, which tend to have large populations, had a higher correlation coefficient between infection rate and poverty rate. We also observe a significant high positive correlation between the infection rate and the number of cumulative cases in Republican counties that had not imposed a face covering order.

## 1 Introduction

The Coronavirus Disease 2019 (COVID-19) is an infectious disease caused by a newly discovered severe acute respiratory syndrome called coronavirus 2 (SARS-SoV-2) (WHO 2021). By March 15, 2021, over 120 million people have been infected with COVID-19, and more than 2.6 million people lost their lives to the virus. The United States is one of the countries most affected by the COVID-19 pandemic. As the second most populous state in the United States, and a state with a mix of rural and urban counties, and both Republican-leaning and Democrat-leaning counties, Texas provides important insights into the impact of various factors on COVID-19 cases. In this paper, we fit Auto Regressive Integrated Moving Average (ARIMA) or seasonal ARIMA models to daily confirmed cases of COVID-19 for all 254 counties in Texas, and cluster the model parameters (*p, d, q*) while accounting for population, political affiliations, face covering order requirements, infection rate, and poverty rate at the county level.

Dehesh, Mardani-Fard and Dehesh (2020) applied ARIMA models for confirmed cases in different countries. Singh et al., (2020) analyzed cumulative cases for the top 15 affected countries using an ARIMA model. Elgar, Stefaniak and Wohl (2020) found the relation between COVID-19 fatalities and social factors including confidence in governments, capital and income inequality for 84 countries. To the best of our knowledge, there has been no analysis about COVID-19 cases at the county level nor including county-level political affiliations. Our aim is to bridge this gap.

Autoregressive Integrated Moving Average (ARIMA) and seasonal ARIMA models were introduced in Box and Jenkins (1970) and explained thoroughly in Box et al., (2015). These models have become widely used in time series analysis. The ARIMA model includes other models as special cases including Moving Average (MA), Autoregressive (AR), and Autoregressive Moving Average models. ARIMA models have been investigated in various applications, such as macroeconomics, finance, environmental science and demography. Further, time series analysis has been frequently used to better understand infectious diseases. Choi and Thacker (1981) evaluated influenza mortality and pneumonia in 1962-1979, while Chang et al., (2004) studied the impact of Severe Acute Respiratory Syndrome (SARS) outbreak in 2003 on medical service utilization. Further, Kane et al., (2014) used an ARIMA model to predict the Highly Pathogenic Avian Influenza (H5N1) outbreak in Egypt.

## 2 Methods

### 2.1 Data Sources

The number of COVID-19 daily confirmed cases for 254 counties in Texas, United States, was obtained from the Texas Department of State Health Services for the period from March 4, 2020 to March 15, 2021 (DSHS 2020). The data on county-level estimated 2019 population is from the Texas Demographic Center (TDC 2019). The 2016 U.S. presidential election results in Texas at the county level is from Harvard Dataverse (Harvard Dataverse 2016). We did not consider the 2020 presidential election results since the COVID-19 pandemic could have affected those results. On July 3, 2020, Texas Governor Greg Abbott signed an executive order (GA-19) to require face covering in public; however, 70 counties in Texas did not enforce the executive order. We label those 70 counties ‘No Face Covering Order’, and label the remaining counties ‘Face Covering Order’. Data regarding the poverty rate for each county in Texas in 2019 is from the 2019 U.S. Census Bureau Small Area Income & Poverty Estimates (SAIPE 2019). In this paper, to calculate the infection rate for each county, we divide that county’s cumulative cases on March 15, 2021 by its population, which *differs* from CDC’s infection rate. We had to adopt a different definition because CDC calculates the infection rate by dividing confirmed cases by total tested cases, but the data about the total tested cases was not available to us.

### 2.2 ARIMA-Model-Based Time Series Analysis

Autoregressive integrated moving average process (ARIMA), first proposed by Box and Jenkins (1970), has been used for fitting time series models in a wide range of areas. In this paper, we will follow the notations in Hyndman and Athanasopoulos (2018) and Brockwell and Davis (2016). ARIMA(*p, d, q*) can be expressed as

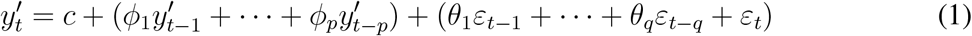

where we use the following notations:

*p* : the order of autoregression (AR).

*d* : the degree of differencing.

*q* : the order of moving average (MA).

*y*_*t*_ : confirmed cases in day *t* in a county.

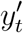: the differenced value of *y*_*t*_.

*c* : the mean of changes between two consecutive days.

*ε*_*t*_ : white noise with mean zero and variance *σ*^2^.

*ϕ* : the parameters of AR part.

*θ* : the parameters of MA part.

Eq. (1) can also be represented in backshift form:

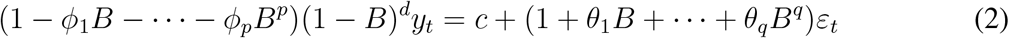

where *B* is the backshift operator. We fit ARIMA or seasonal ARIMA models for each of the 254 Texas counties using the Hyndman-Khandakar algorithm (Hyndman and Khandakar 2008), and use the R software package contributed by Hyndman and Khandakar:

1. Plot the daily confirmed cases time series to identify any usual observation such as obvious backlog reported in the Texas Department of State Health Services system.
2. Perform a Box-Cox transformation if the time series has a pattern of changing variance. In this paper, we apply the natural logarithm to (*y*_*t*_ + 1) for the time series.
3. Difference the data until it achieves stationarity by conducting the Kwiatkowski-Phillips-Schmidt-Shin (KPSS) test (Kwiatkowski et al., 1992).
4. Select the AR parameter *p* and the MA parameter *q* based on plots of autocorrelation function (ACF) and partial autocorrelation function (PACF). Then, select *p, q, ϕ*_*p*_ and *θ*_*q*_ for candidate models with lower Corrected Akaike’s Information Criterion (AICc). In this part, the R function auto.arima based on AICc will be applied. We set the maximum value of *p* + *q* + *P* + *Q* to 10, force it to search over all models without stepwise selection, and allow it to search seasonal models. Then, we record the 10 models with lowest AICc.
5. Conduct Ljung-Box tests for the residuals of the recorded 10 models, and keep the models that fail to reject the null hypothesis at a significance level of 0.05 as our candidate models. We label the candidate model with the lowest AICc as Rank1, the second lowest AICc as Rank2, etc.

For counties that cannot be successfully fitted to ARIMA models using this algorithm (i.e., no model passes the Ljung-Box test), we will consider the seasonality. The seasonal ARIMA(*p, d, q*)(*P, D, Q*)_*m*_ with period *m* can be represented as:

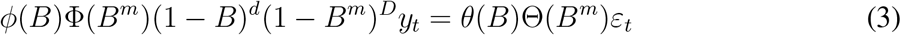

where *ϕ*(*B*) = 1*−ϕ*_1_*B −…−ϕ*_*p*_*B*^*p*^, Φ(*B*) = 1*−*Φ_1_*B −…−*Φ_*P*_ *B*^*P*^, *θ*(*B*) = 1*−θ*_1_*B −…−θ*_*q*_*B*^*q*^, and Θ(*B*) = 1 *−* Θ_1_*B − … −* Θ_*Q*_*B*^*Q*^. The following example illustrates our models for Dallas County, Texas. In Figure 1, we do not observe any unusual data point related to input error or backlog, and we plot *log*(*y*_*t*_ + 1) for further analysis. Based on Figure 2 showing the differencing of data with taking the logarithm, and conducting the KPSS test, taking one differencing (*diff* (*log*(*y*_*t*_ + 1))) achieves stationarity (second panel in Figure 2). The ACF and PACF plots of *diff* (*log*(*y*_*t*_ + 1)) are shown in Figure 3.

**Figure 1:**
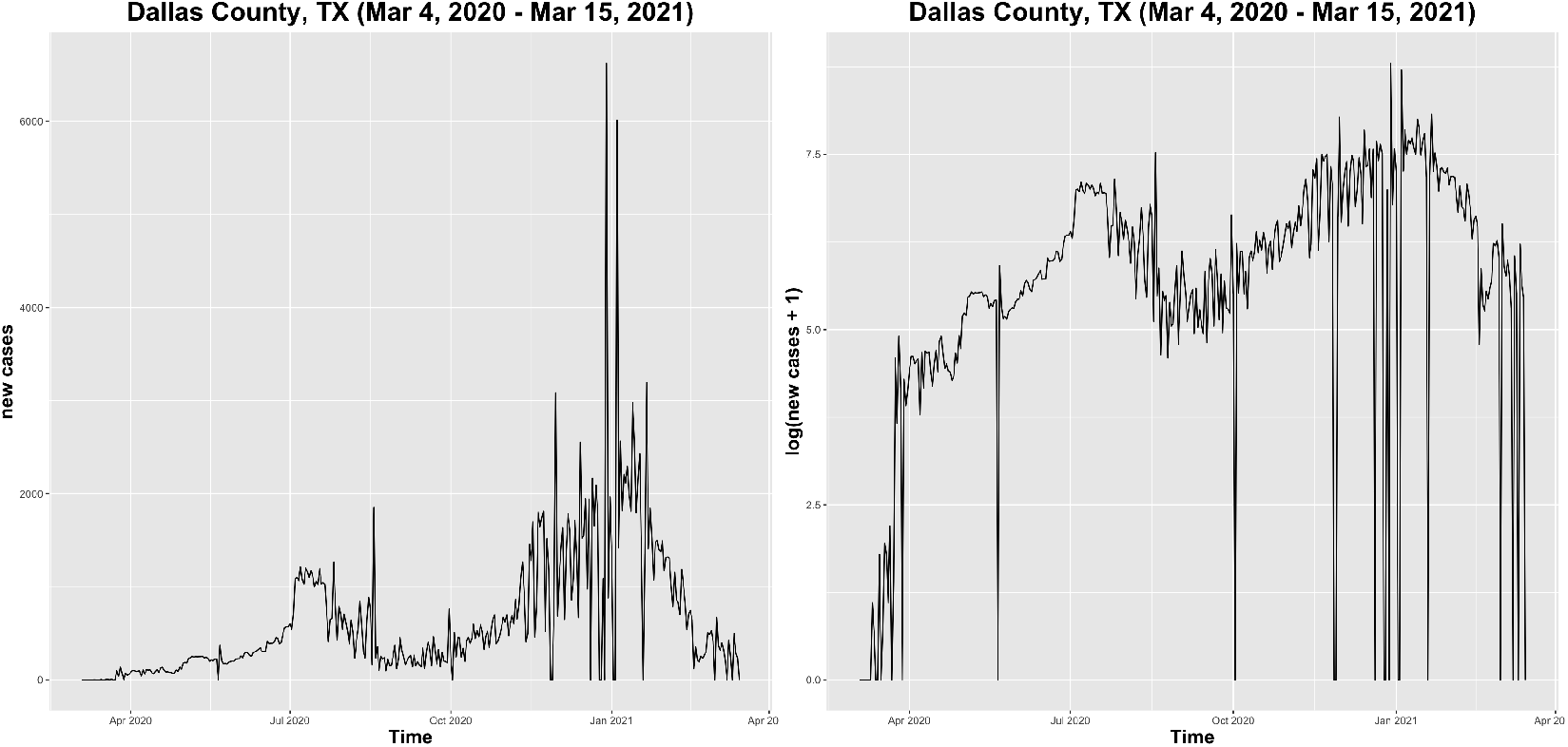
New cases (left) and log(new cases + 1) (right) in Dallas County, TX, USA

**Figure 2:**
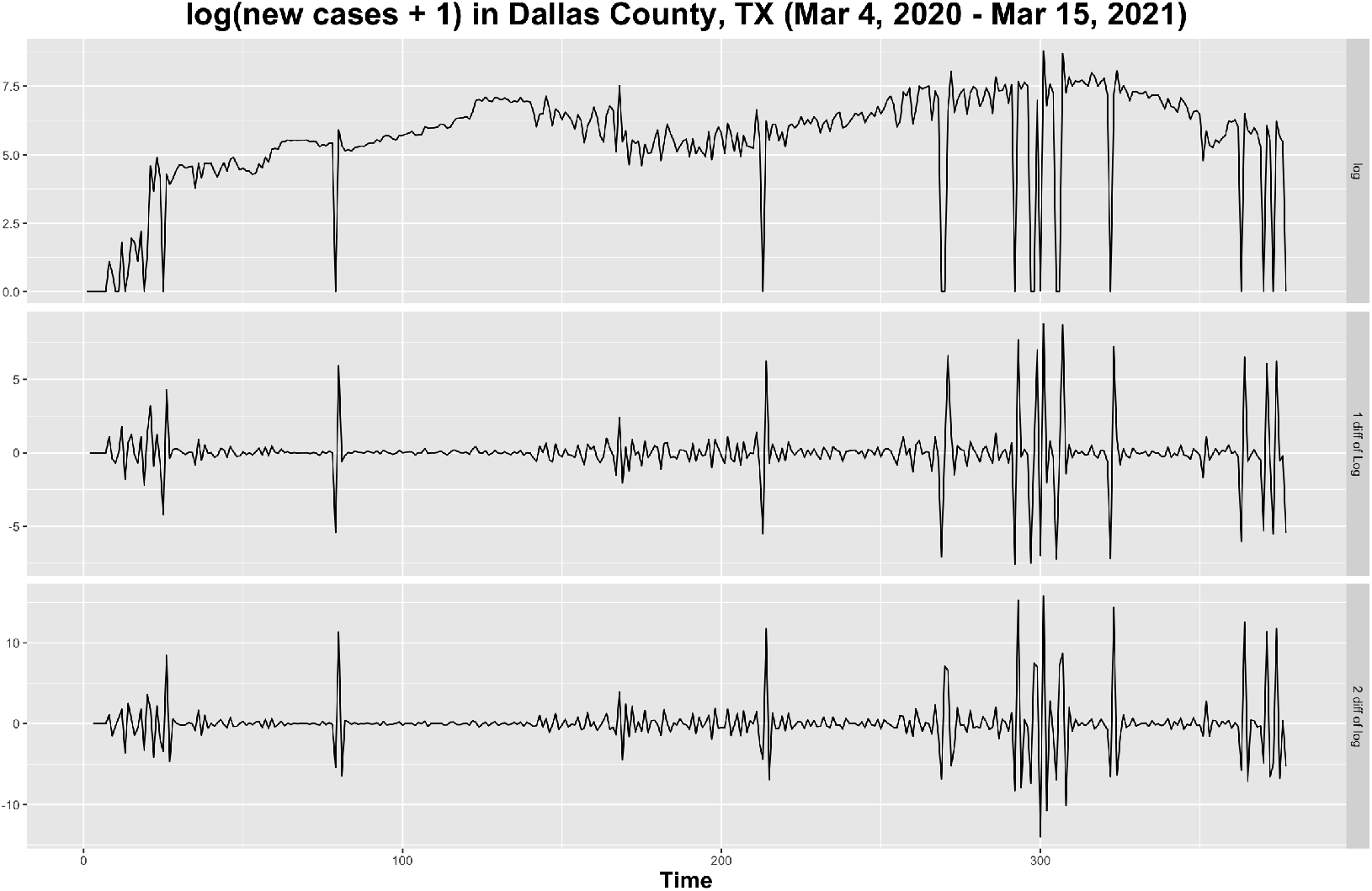
Differencing of log(new cases + 1) in Dallas County, TX, USA

**Figure 3:**
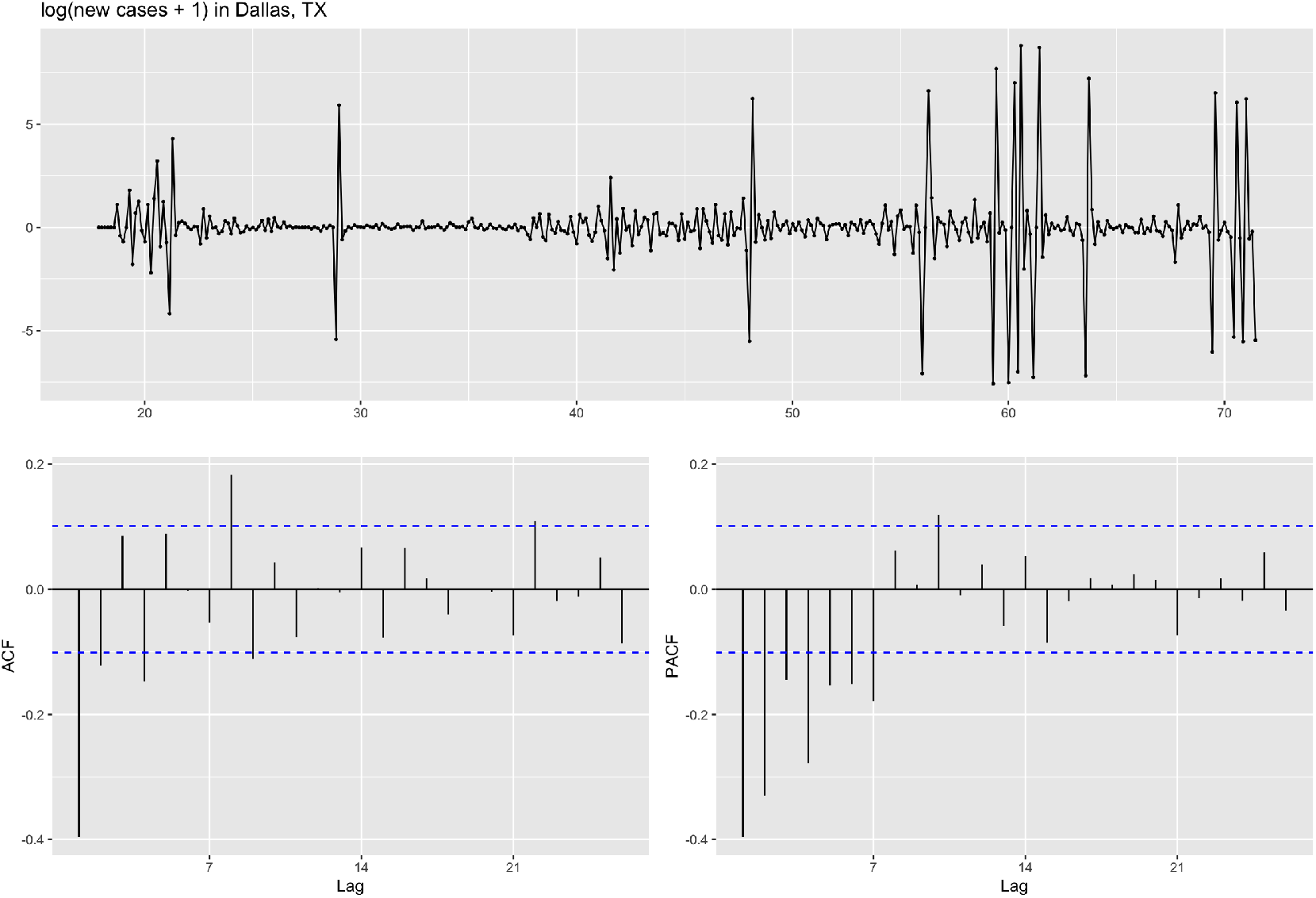
ACF and PACF of log(new cases + 1) in Dallas County, TX, USA

291 models for Dallas are fitted, and 10 models with lowest AICc are recorded. Next, we perform Ljung-Box tests for those 10 models, and all of them fail to reject the null hypothesis at a significance level of 0.05. Thus, Dallas County, TX has 10 candidate models for *log*(*y*_*t*_ + 1) and the result is shown in Table 1. L-B test is shorthand for Ljung-Box test.

**Table 1:** Summary of ten candidate models for Dallas County, TX, USA

| Rank | Model with logarithm | AICc | L-B test (p-value) | MAE | RMSE | MASE |
| --- | --- | --- | --- | --- | --- | --- |
| 1 | ARIMA(5,1,5) | 1324.582 | 0.5841 | 0.7315688 | 1.361112 | 0.8279928 |
| 2 | ARIMA(5,1,4) | 1327.511 | 0.183 | 0.7253109 | 1.370882 | 0.8209101 |
| 3 | ARIMA(4,1,5) | 1328.323 | 0.2303 | 0.7207021 | 1.372387 | 0.8156939 |
| 4 | ARIMA(2,1,5) | 1329.513 | 0.0531 | 0.7295895 | 1.381834 | 0.8257527 |
| 5 | ARIMA(4,1,2)(1,0,1)[7] | 1329.851 | 0.05868 | 0.7174613 | 1.379131 | 0.8120258 |
| 6 | ARIMA(4,1,2)(1,0,0)[7] | 1329.855 | 0.1126 | 0.7101057 | 1.383177 | 0.8037008 |
| 7 | ARIMA(4,1,5)(0,0,1)[7] | 1329.903 | 0.19 | 0.7201692 | 1.371393 | 0.8150907 |
| 8 | ARIMA(4,1,2)(0,0,1)[7] | 1330.112 | 0.1077 | 0.71024 | 1.383668 | 0.8038528 |
| 9 | ARIMA(2,1,5)(0,0,1)[7] | 1330.277 | 0.1124 | 0.7204406 | 1.379949 | 0.8153979 |
| 10 | ARIMA(4,1,3) | 1330.401 | 0.1242 | 0.7158871 | 1.384165 | 0.8102442 |

### 2.3 Clustering Method

We perform ARIMA or seasonal ARIMA models for each of the 254 counties in Texas in a way similar to our approach for Dallas County. Next, we cluster the parameters (*p, d, q*) of the candidate models for all counties in Texas, and analyze counties in the same cluster to identify whether they have similar characteristics regarding population size, cumulative cases, political affiliations, poverty rate, infection rate, or face covering requirements.

The approach to cluster non-seasonal ARIMA components (*p, d, q*) for all counties is as follows:

1. List non-seasonal ARIMA components (*p, d, q*) of Rank1 models for all counties, and those (*p, d, q*) of Rank1 models are grouped into (for this data set) 38 unique clusters.
2. Attempt to combine the clusters that consist of only one county into other existing clusters (by using models of lower rank instead of Rank1 models, if those (*p, d, q*) already appear in the list of existing clusters) to reduce the total number of clusters. We observe that 11 (*p, d, q*) clusters only contain one county. For example, the (2, 0, 5) cluster only includes Hidalgo County. Hidalgo’s Rank2 (*p, d, q*) is (1, 0, 2) which cannot be combined since (1, 0, 2) does not appear in the list of existing clusters. Then, we check Hidalgo’s Rank3 (*p, d, q*), and the Rank3 (1, 0, 3) can be combined into the existing cluster (1, 0, 3). Thus, after conducting the step of combining Hidalgo, the cluster (2, 0, 5) is removed, and the total number of unique clusters is reduced to 37.
3. For the situation of cluster (*p, d, q*) including only one county that cannot be combined to other existing clusters, we would keep it. For instance, cluster (5, 0, 3) only contains Shelby County, and other Shelby’s Ranks (*p, d, q*) cannot be combined into any existing clusters. We would keep cluster (5, 0, 3) in the result.
4. Apply similar procedures to combine (*p, d, q*) clusters that consist of only two counties.
5. Apply the similar procedures to combine (*p, d, q*) clusters that consist of only three counties.

## 3 Results

Among the 254 counties in Texas, 223 of them have at least one candidate ARIMA or seasonal ARIMA model, and the remaining 31 counties cannot be fitted to any model that fails to reject the null hypothesis of Ljung-Box tests at a significance level of 0.05. Only one county among the 31 is Democrat-leaning, and the remaining 30 counties are Republican-leaning. In this paper, we focus on the analysis of the 223 counties that have candidate models.

The 223 counties are grouped into 23 clusters in terms of (*p, d, q*) components. The results are summarized in Table 2, and larger values are highlighted into darker color. The most common clusters are (0, 1, 1) (17.04% of counties) and (2, 1, 3) (9.42% of counties). (Patterns for time series in those clusters are illustrated in Figure 7 below.) The geographic plots for clusters of all 223 counties, 26 Democratic counties, and 197 Republican counties are presented in Figures 4, 5 and 6, where the circle size in the plots represents the estimated 2019 Texas population at the county level.

**Table 2:**
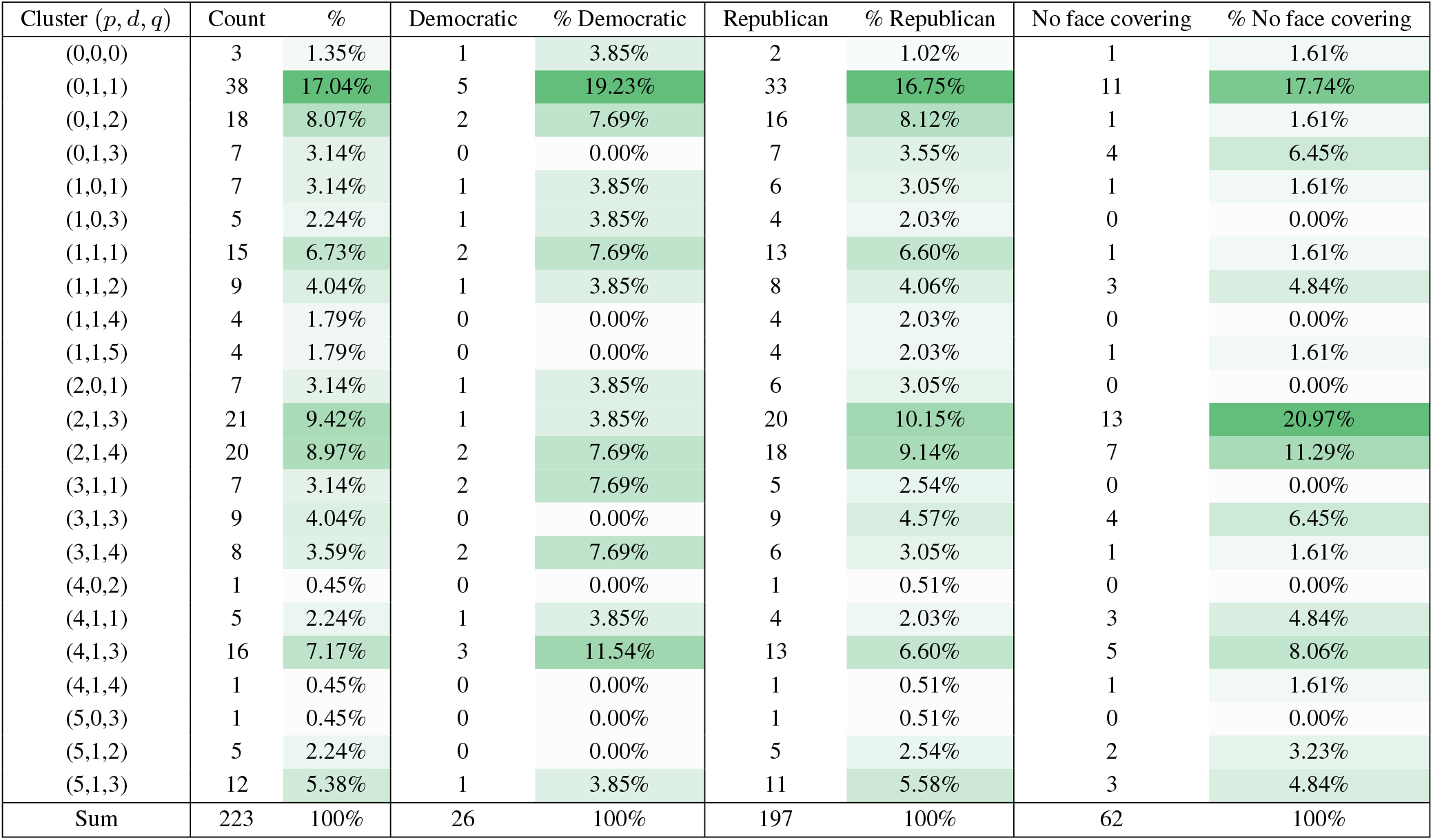
Summary of clusters for 223 counties in Texas

| Cluster ($p, d, q$) | Count | % | Democratic | % Democratic | Republican | % Republican | No face covering | % No face covering |
| --- | --- | --- | --- | --- | --- | --- | --- | --- |
| (0,0,0) | 3 | 1.35% | 1 | 3.85% | 2 | 1.02% | 1 | 1.61% |
| (0,1,1) | 38 | 17.04% | 5 | 19.23% | 33 | 16.75% | 11 | 17.74% |
| (0,1,2) | 18 | 8.07% | 2 | 7.69% | 16 | 8.12% | 1 | 1.61% |
| (0,1,3) | 7 | 3.14% | 0 | 0.00% | 7 | 3.55% | 4 | 6.45% |
| (1,0,1) | 7 | 3.14% | 1 | 3.85% | 6 | 3.05% | 1 | 1.61% |
| (1,0,3) | 5 | 2.24% | 1 | 3.85% | 4 | 2.03% | 0 | 0.00% |
| (1,1,1) | 15 | 6.73% | 2 | 7.69% | 13 | 6.60% | 1 | 1.61% |
| (1,1,2) | 9 | 4.04% | 1 | 3.85% | 8 | 4.06% | 3 | 4.84% |
| (1,1,4) | 4 | 1.79% | 0 | 0.00% | 4 | 2.03% | 0 | 0.00% |
| (1,1,5) | 4 | 1.79% | 0 | 0.00% | 4 | 2.03% | 1 | 1.61% |
| (2,0,1) | 7 | 3.14% | 1 | 3.85% | 6 | 3.05% | 0 | 0.00% |
| (2,1,3) | 21 | 9.42% | 1 | 3.85% | 20 | 10.15% | 13 | 20.97% |
| (2,1,4) | 20 | 8.97% | 2 | 7.69% | 18 | 9.14% | 7 | 11.29% |
| (3,1,1) | 7 | 3.14% | 2 | 7.69% | 5 | 2.54% | 0 | 0.00% |
| (3,1,3) | 9 | 4.04% | 0 | 0.00% | 9 | 4.57% | 4 | 6.45% |
| (3,1,4) | 8 | 3.59% | 2 | 7.69% | 6 | 3.05% | 1 | 1.61% |
| (4,0,2) | 1 | 0.45% | 0 | 0.00% | 1 | 0.51% | 0 | 0.00% |
| (4,1,1) | 5 | 2.24% | 1 | 3.85% | 4 | 2.03% | 3 | 4.84% |
| (4,1,3) | 16 | 7.17% | 3 | 11.54% | 13 | 6.60% | 5 | 8.06% |
| (4,1,4) | 1 | 0.45% | 0 | 0.00% | 1 | 0.51% | 1 | 1.61% |
| (5,0,3) | 1 | 0.45% | 0 | 0.00% | 1 | 0.51% | 0 | 0.00% |
| (5,1,2) | 5 | 2.24% | 0 | 0.00% | 5 | 2.54% | 2 | 3.23% |
| (5,1,3) | 12 | 5.38% | 1 | 3.85% | 11 | 5.58% | 3 | 4.84% |
| Sum | 223 | 100% | 26 | 100% | 197 | 100% | 62 | 100% |

**Figure 4:**
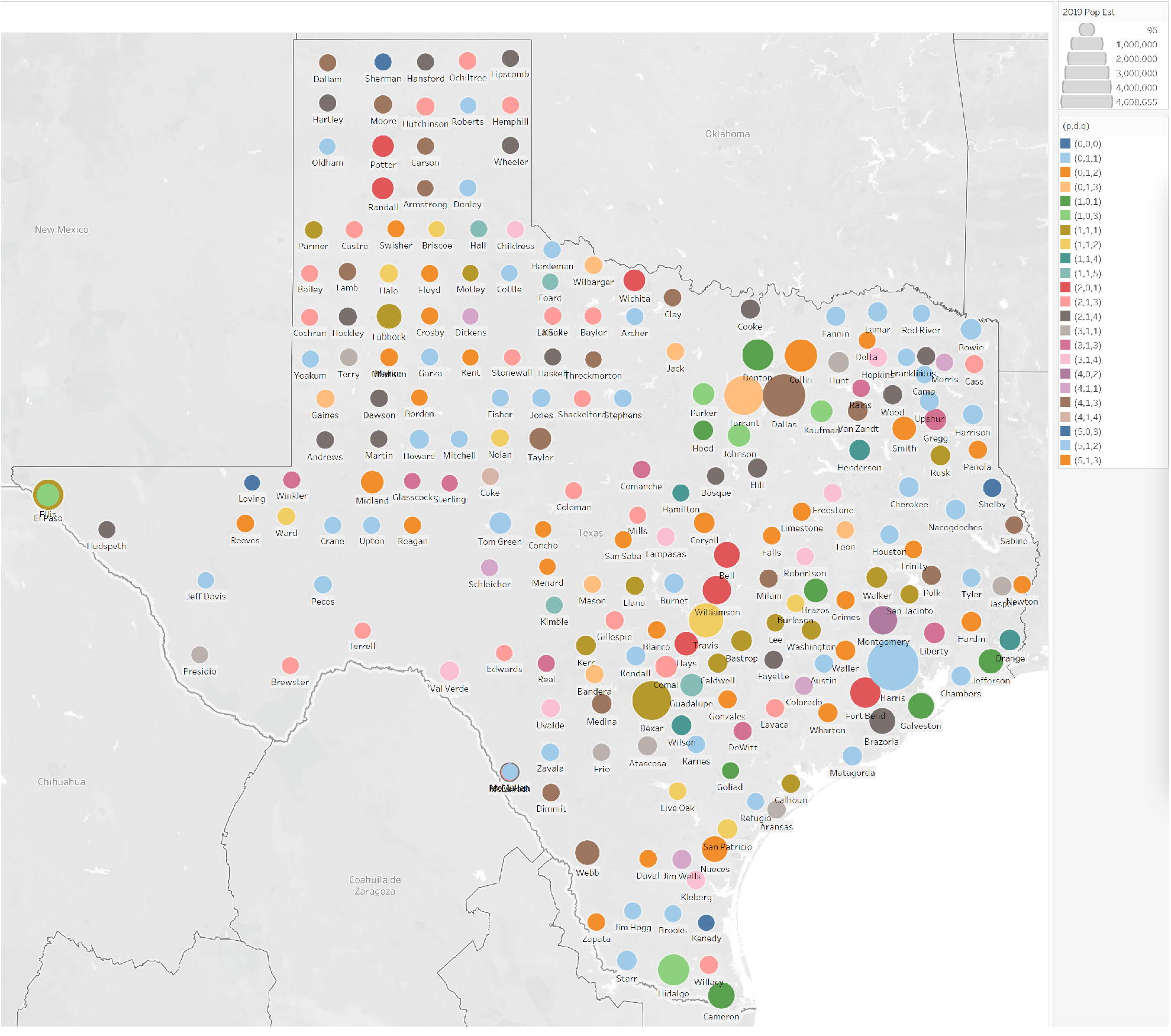
Clusters of all counties in Texas

**Figure 5:**
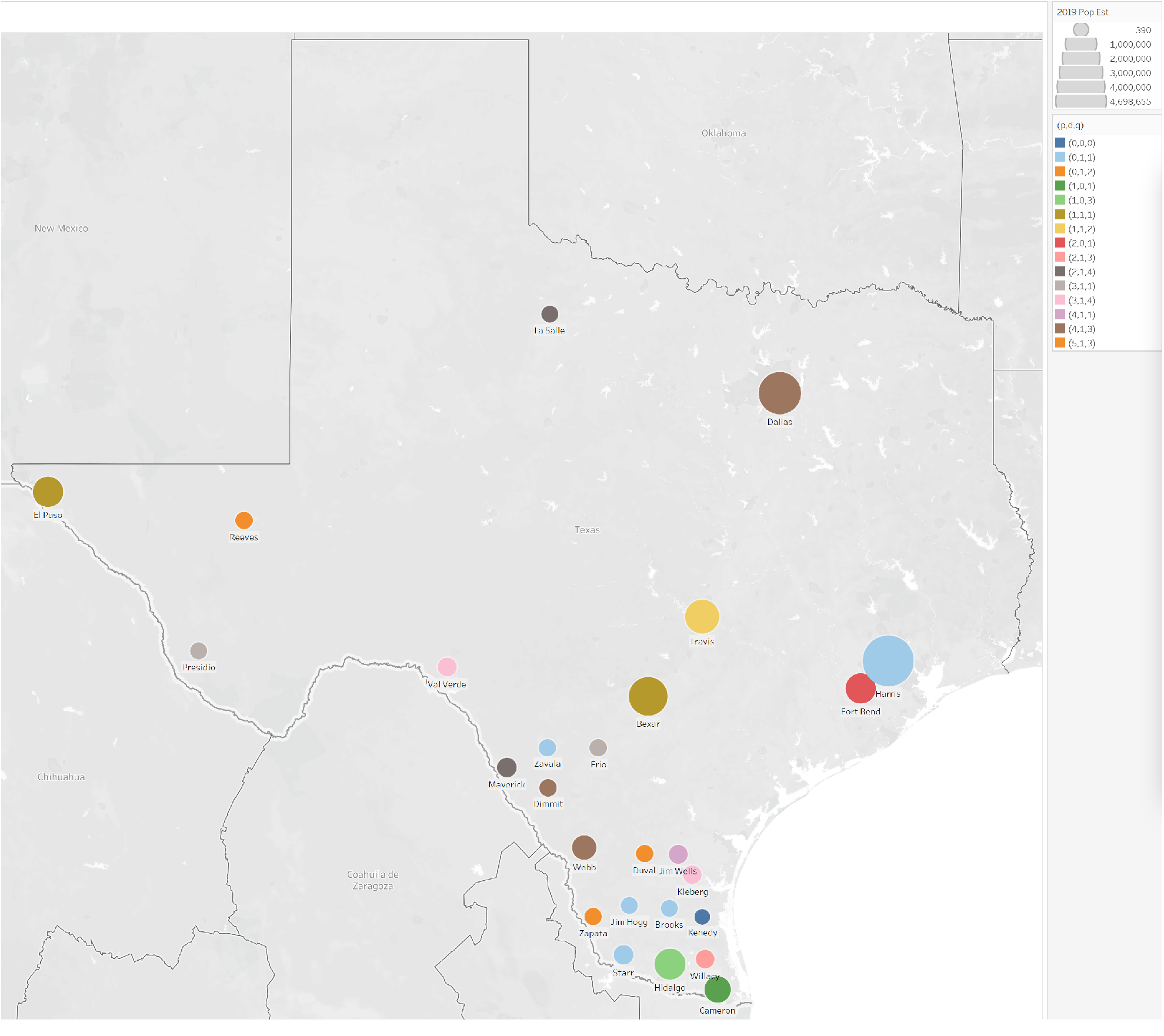
Clusters of Democratic counties in Texas

**Figure 6:**
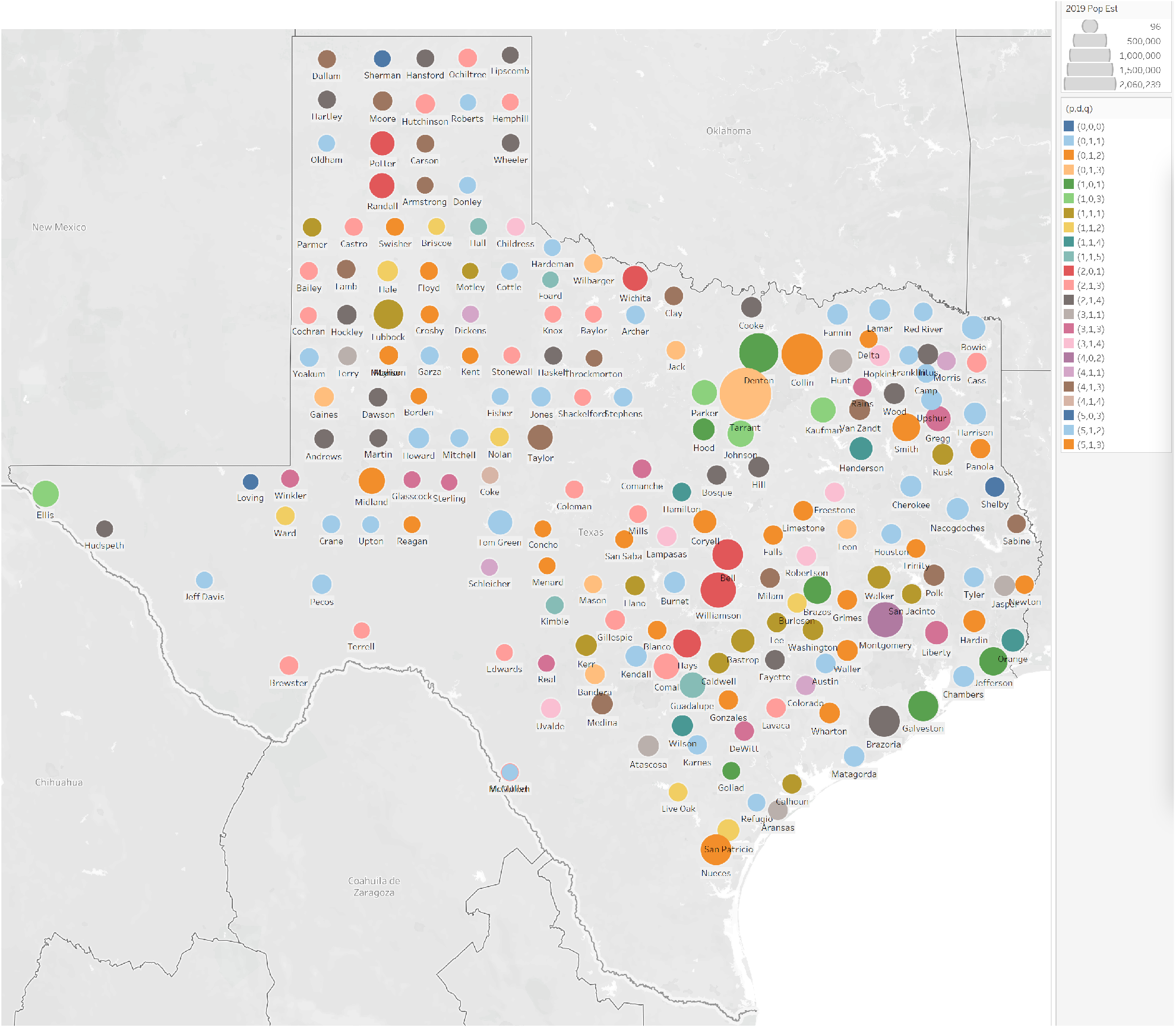
Clusters of Republican counties in Texas

**Figure 7:**
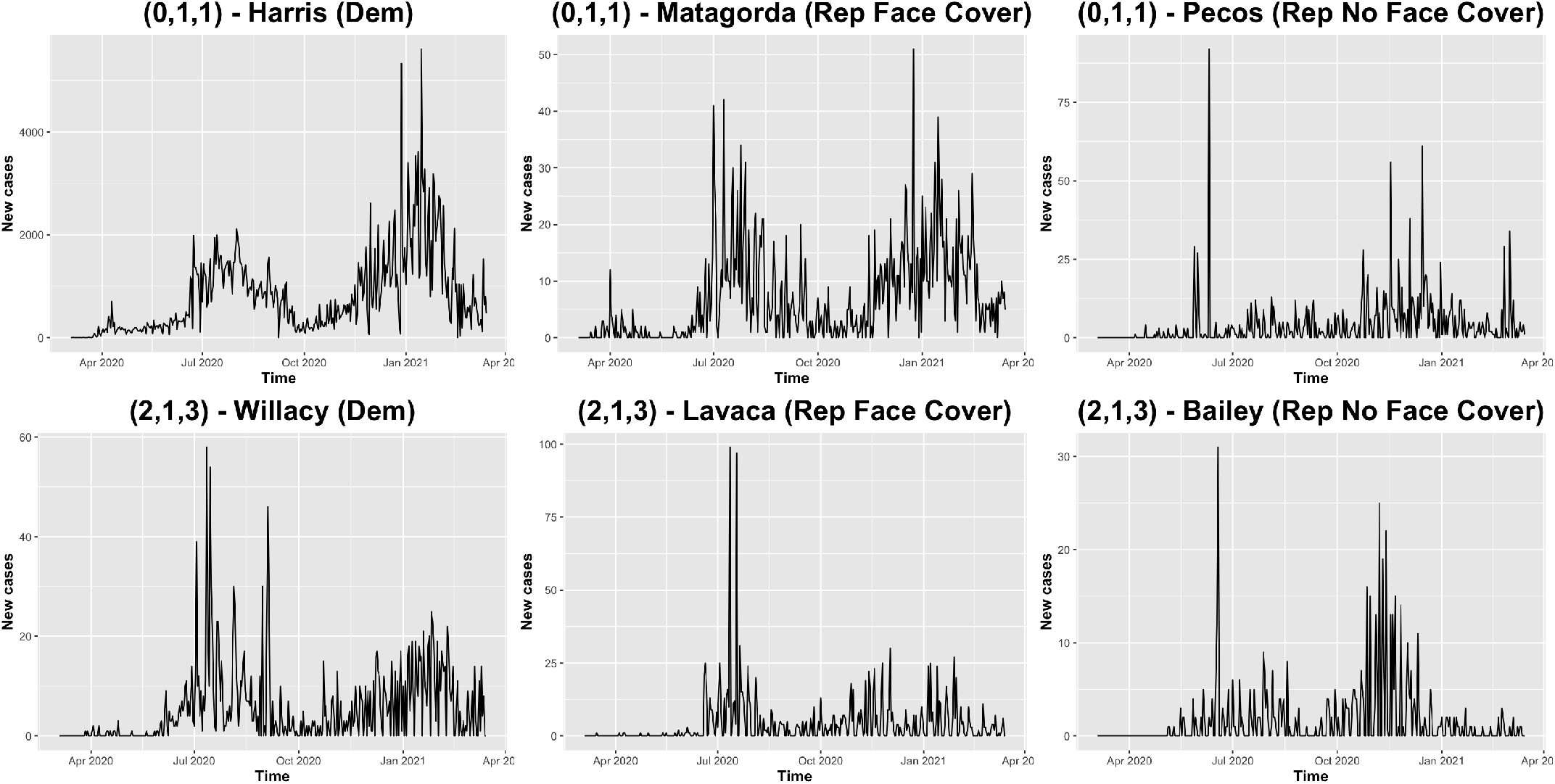
Examples of time series in the most common (*p, d, q*) clusters for counties in different scenarios

**Figure 8:**
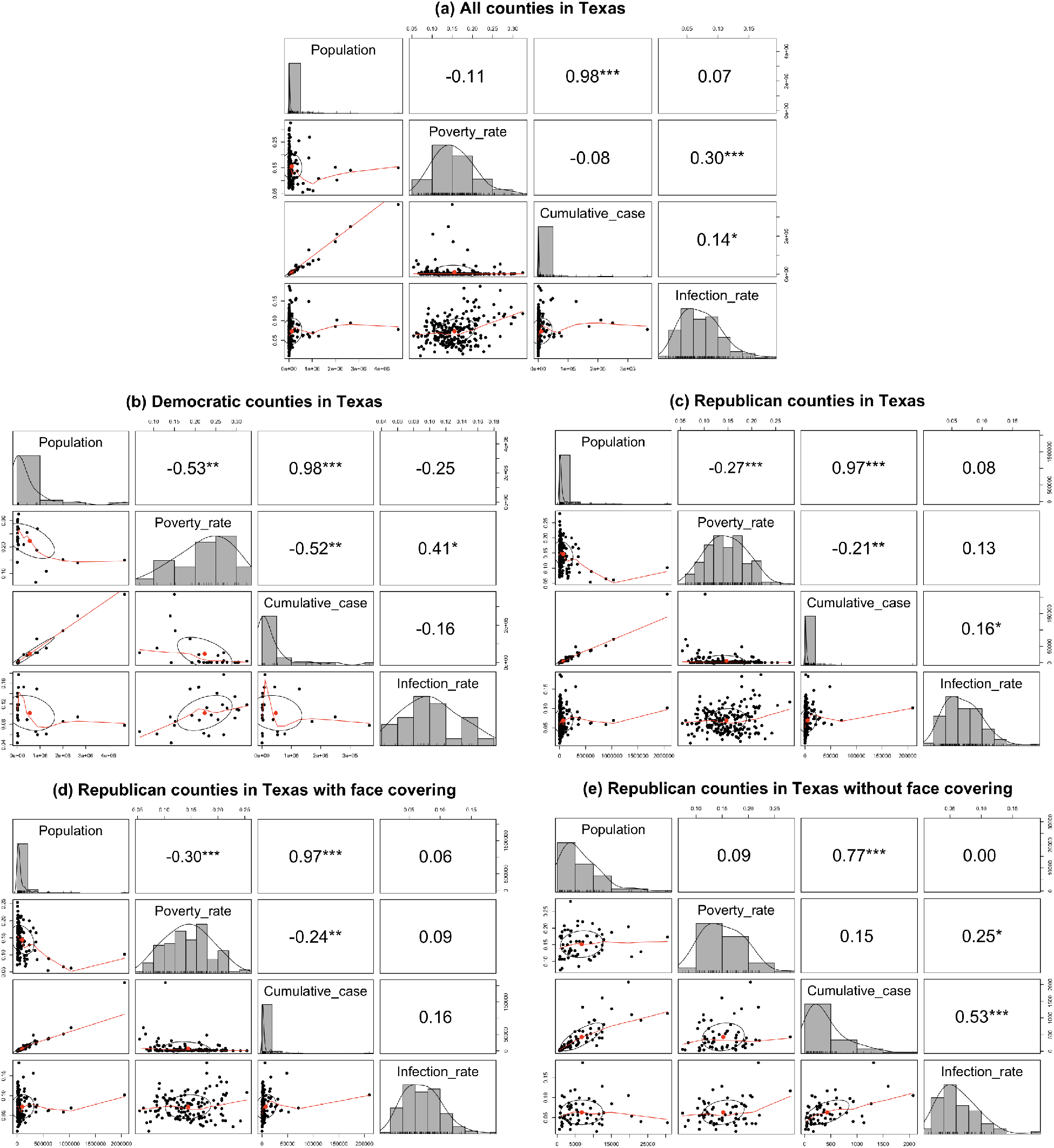
Correlation matrix

Nearly 50% of 223 counties are clustered into (0, 1, 1), (2, 1, 3), (2, 1, 4), (0, 1, 2) and (4, 1, 3). Democratic counties are grouped into 15 unique (*p, d, q*) clusters, and no cluster except (0, 1, 1) contains a lot more counties than other clusters. For Republican counties, 36.04% counties are clustered into (0, 1, 1), (2, 1, 3) and (2, 1, 4). For the counties that did not impose a face covering order, which are all Republican counties with relatively smaller population size, 50% of them are clustered into (2, 1, 3), (0, 1, 1) and (2, 1, 4).

Figure 7 shows actual examples of time series in the most common clusters (0, 1, 1) and (2, 1, 3) distinguishing by political affiliations and face covering order, where “Dem” and “Rep” are abbreviations of Democratic and Republican respectively. The patterns of time series of counties with face covering order clustered in (0, 1, 1) and (2, 1, 3) differ, that daily confirmed cases of (2, 1, 3) has a large spike (in this case in July 2020) and drops *suddenly* afterwards; however, daily confirmed cases of (0, 1, 1) *gradually* decreases after the emergence of a spike (in this case in July 2020), and have a large spike (in January 2021) afterwards.

We take political affiliations and face covering orders into account, and compute Pearson’s correlation coefficients between population, cumulative cases, poverty rate and infection rate at the county level in each of the following cases: all counties, Democratic counties, Republican counties, Republican counties with face covering order, and Republican counties without face covering order. Note that all Democratic counties in Texas imposed face a covering order, so we do not need to distinguish between with and without face covering order there. Then, we compute correlation coefficients between numerical variables for scenarios with and without face covering order in Republican counties. The correlation matrix and significant Pearson’s correlation coefficients are provided in Figure 6 and Table 3 respectively.

**Table 3:** Significant Pearson’s correlation coefficients

| **All counties** |  | cor | p-value | **Democratic counties** |  | cor | p-value |
| --- | --- | --- | --- | --- | --- | --- | --- |
| Population | Cumulative cases | 0.98 | 0.00 | Population | Cumulative cases | 0.98 | 0.00 |
| Poverty rate | Infection rate | 0.30 | 0.00 | Poverty rate | Infection rate | 0.41 | 0.04 |
| Cumulative cases | Infection rate | 0.14 | 0.04 | Poverty rate | Cumulative cases | -0.52 | 0.01 |
|  |  |  |  | Poverty rate | Population | -0.53 | 0.01 |
| **Republican counties** |  | cor | p-value | **Republican counties with face covering** |  | cor | p-value |
| Population | Cumulative cases | 0.97 | 0.00 | Population | Cumulative cases | 0.97 | 0.00 |
| Cumulative cases | Infection rate | 0.16 | 0.02 | Infection rate | Cumulative cases | 0.16 | 0.06 |
| Poverty rate | Infection rate | 0.13 | 0.07 | Poverty rate | Cumulative case | -0.24 | 0.01 |
| Poverty rate | Cumulative cases | -0.21 | 0.00 | Poverty rate | Population | -0.30 | 0.00 |
| Poverty rate | Population | -0.27 | 0.00 |  |  |  |  |
| **Republican counties without face covering** |  | cor | p-value |  |  |  |  |
| Population | Cumulative cases | 0.77 | 0.00 |  |  |  |  |
| Poverty rate | Infection rate | 0.25 | 0.05 |  |  |  |  |
| Cumulative cases | Infection rate | 0.53 | 0.00 |  |  |  |  |

Population and cumulative cases are highly positively correlated in all cases significantly, which matches our intuition. In cases considering Democratic counties, Republican counties, and Republican counties with face covering, population and poverty rate are significantly negatively correlated. Counties with a larger population tend to have a smaller poverty rate no matter what political affiliations; however, Democratic counties with a larger population have a far smaller poverty rate generally. In addition, poverty rate and infection rate are significantly positively correlated in cases considering Democratic counties and Republican counties without a face covering order; however, the correlation between poverty rate and infection rate is not significant in Republican counties with a face covering order. In the case of Republican counties without a face covering order, infection rate and cumulative cases have a high positive correlation coefficient of 0.53. The population of these counties is relatively small, namely these counties have fewer than 30,000 residents.

The box plots of county population, cumulative cases, infection rate, poverty rate for all the (*p, d, q*) clusters of the 223 counties are provided in Figure 9. In general, counties with larger medians of population and cumulative cases are clustered in (1, 0, 1) and (2, 0, 1). In addition, counties with lower median of infection rate are clustered in (0, 0, 0), and counties with lower median of poverty rate are clustered in (1, 0, 3).

**Figure 9:**
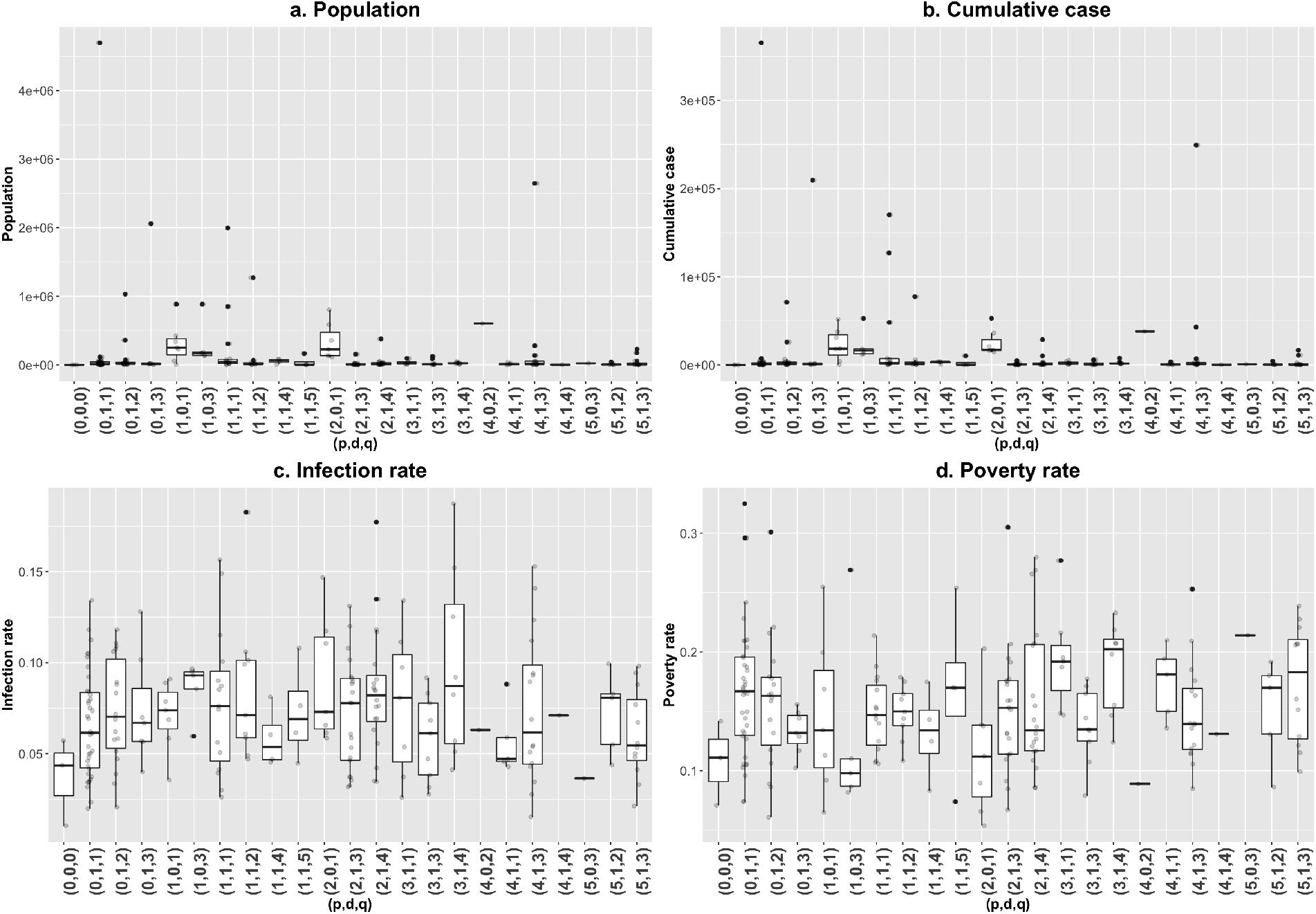
Box plots of clusters (*p, d, q*)

We also analyze thoroughly the impact of political affiliations and face covering orders on our clustering results. Clusters of (*p, d, q*) parameters for Democratic and Republican counties, and distinguishing between whether a face covering order was imposed or not, are presented in Figure 10 and Figure 11, respectively. Since all Democratic counties imposed a face covering order, Figure 11 investigates the face covering order in Republican counties only.

**Figure 10:**
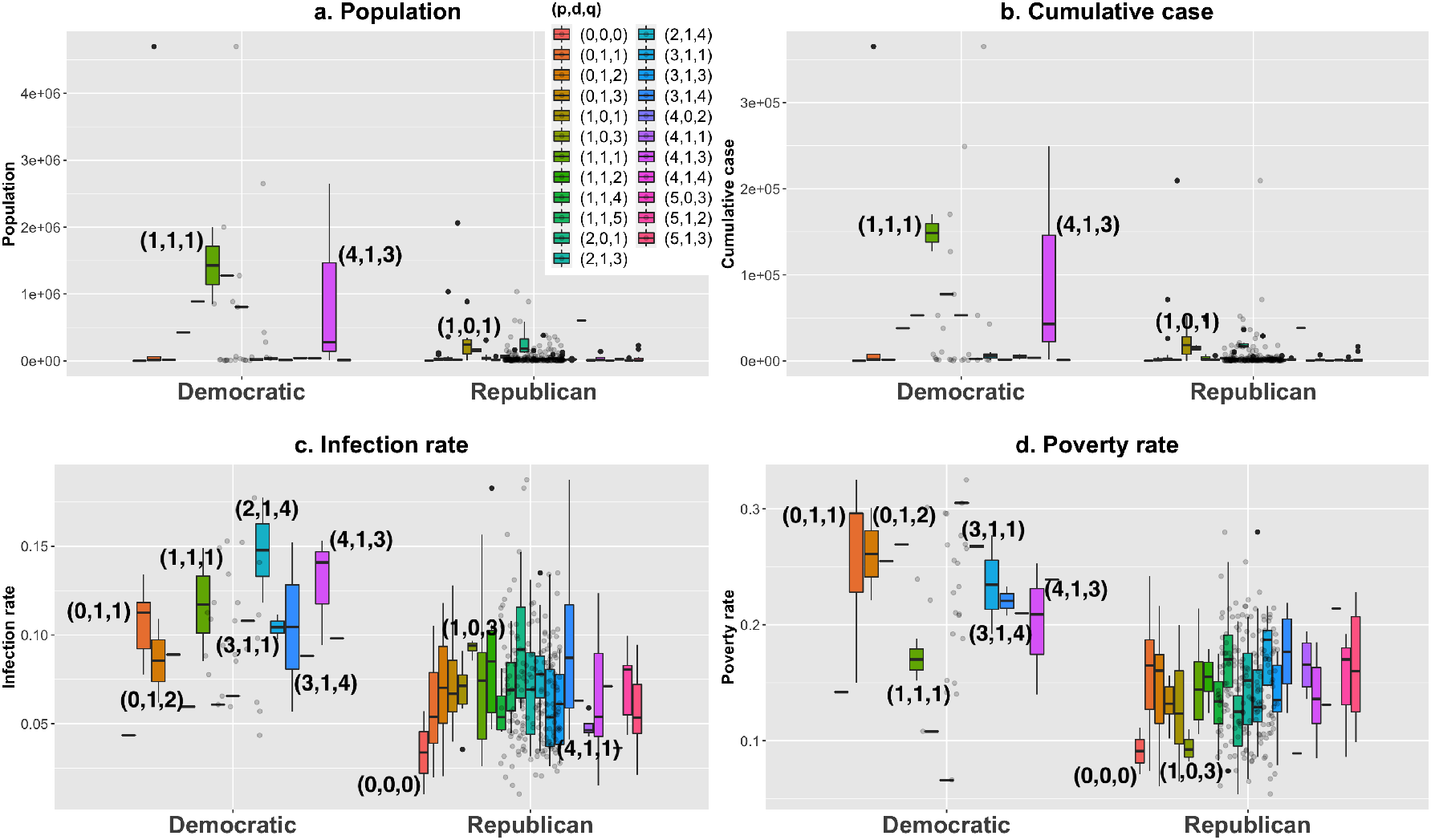
Box plots of (*p, d, q*) varying political affiliations

**Figure 11:**
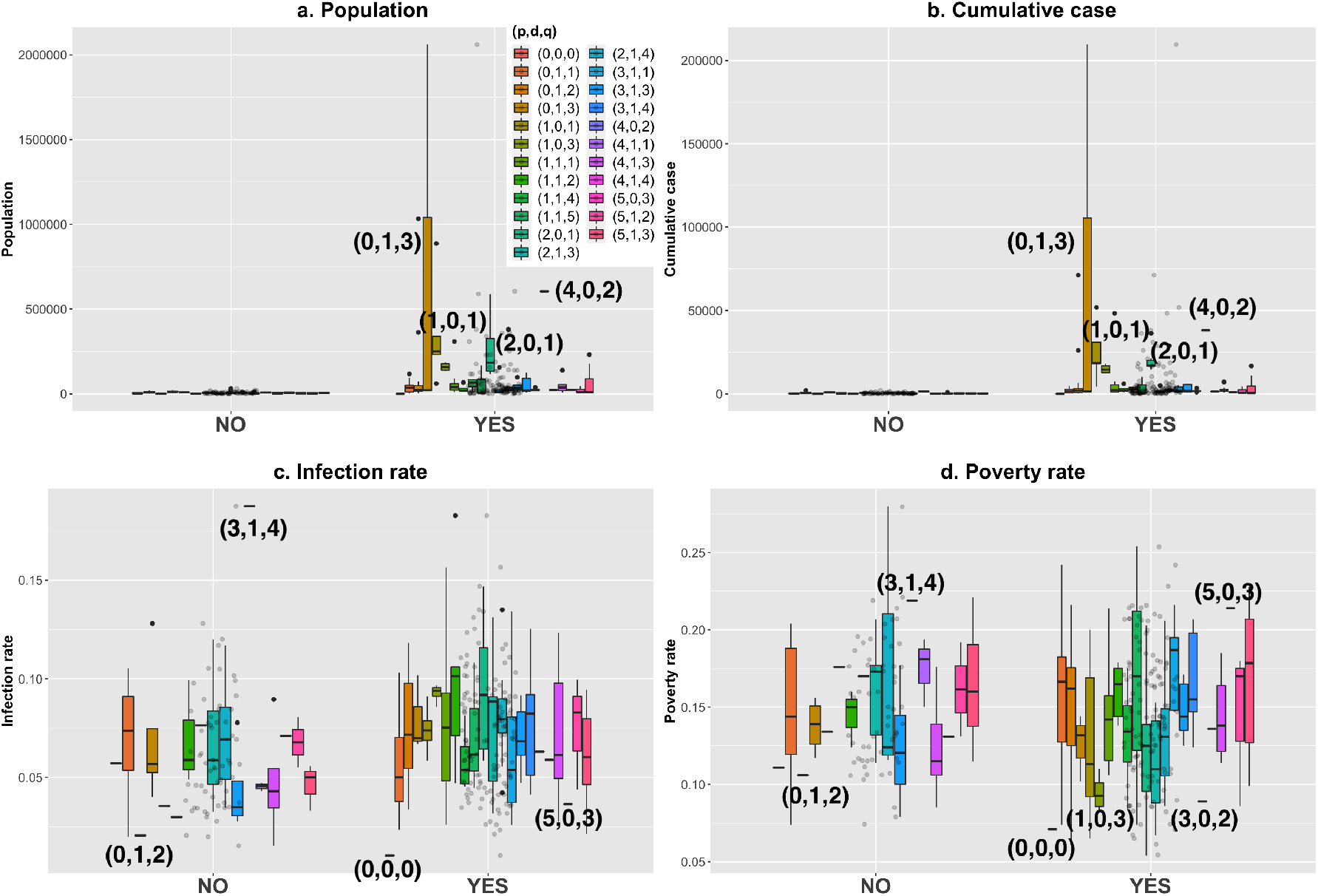
Box plots of (*p, d, q*) varying face covering order in Republican counties

Democratic counties with higher median population and cumulative cases are clustered in (1, 1, 1), and Democratic counties with wider interquartile range of population and cumulative cases are clustered in (4, 1, 3). However, clusters of Republican counties have relatively lower median population and cumulative cases. Regarding the infection rate, clusters of Democratic counties have higher median value, and highest median infection rate for the Democratic counties is clustered in (2, 1, 4). For the Republican counties, the lowest and highest median infection rate are clustered in (0, 0, 0) and (1, 0, 3), respectively. For the poverty rate, clusters of Democratic counties have a wider range of medians compared to the Republican counties. For Republican counties with lower median poverty rate are clustered in (0, 0, 0) and (1, 0, 3).

Regarding the case whether a face covering order was imposed in Republican counties, as mentioned previously, Republican counties that did not impose a face covering order in public have a far smaller population and fewer cumulative cases. The Republican counties that imposed a face covering order have a higher median in population and the cumulative cases are clustered in (4, 0, 2), (1, 0, 1) and (2, 0, 1). For the counties without face covering order, the highest value for both infection rate and poverty rate are clustered in (3, 1, 4). Note that in the case of counties without a face covering order, cluster (3, 1, 4) only includes Childress County. For the Republican counties with a face covering order, cluster (5, 0, 3) has the highest poverty rate and a lower infection rate, and cluster (5, 0, 3) only contains Shelby County.

## 4 Conclusion

The current COVID-19 pandemic has had disastrous consequences for global public health. The United States has seen more than 30 million cumulative positive cases and more than 550, 000 fatalities. Texas is one of the U.S. states most affected by the COVID-19 pandemic. In this paper, we have proposed a time series analysis for all counties in Texas. We have performed ARIMA or seasonal ARIMA models to fit the logarithm of daily confirmed cases for each county, and clustered counties according to the models’ non-seasonal (*p, d, q*) parameters. We consider various cases for the predominant political affiliation and face covering orders for all Texas counties, and take population, infection rate, and poverty rate into consideration. Our analysis has found that, in Texas, clusters of (*p, d, q*) parameters are related to population, infection rate and poverty rate for different combinations of the predominant political affiliation and face covering orders. We also found that the infection rate and poverty rate had significant high positive correlation, with a higher correlation coefficient in Democratic counties than in Republican counties. In Republican counties without a face covering order, the infection rate and the cumulative cases have a significant high positive correlation coefficient, which is not observed in other scenarios. In subsequent research, we intend to analyze how the clusters will change under a new administration after the election of November 2020, different vaccination rates in the counties and the spread of multiple variants of the virus.

## Data Availability

As explained in the data section of the paper, all the data we used is publicly available online and can be retrieved from the links provided below.

https://dshs.texas.gov/coronavirus/AdditionalData.aspx

https://dataverse.harvard.edu/dataset.xhtml?persistentId=doi:10.7910/DVN/VOQCHQ

https://www.census.gov/library/publications/2020/demo/p30-08.html

https://demographics.texas.gov/data/tpepp/estimates/

## Acknowledgements

The authors would like to thank the Hart Center for Engineering Leadership at SMU for supporting the research of the second author through a Lyle Summer Undergraduate Research Fellowship in Summer 2020 and the attendees of the Undergraduate Research Symposium at SMU in October 2020 for insightful comments on preliminary results for this project.

